## Supplementary Appendix for "The prognostic and diagnostic value of intraleukocytic malaria pigment: an individual patient data pooled meta-analysis of 32,000 patients with severe falciparum malaria in Africa and Asia"

September 5, 2022

### Supplementary Tables and Figures

Table S1: Pooled individual patient data set. The median (range) age is shown. $n$ is the total number of patients enrolled in each study.

| Study | Countries | Years | n | Deaths | Age (years) | Coma | PMN counts | PMM counts | Parasite counts |
|---|---|---|---|---|---|---|---|---|---|
| AQ Vietnam [1] | Vietnam | 1991-1996 | 560 | 83 (15%) | 30 (15-79) | 52% | 483 | 301 | 560 |
| SEAQUAMAT [2] | Myanmar, India, Bangladesh, Indonesia | 2003-2005 | 1461 | 270 (19%) | 25 (2-87) | 43% | 1330 | 1329 | 1457 |
| AQUAMAT [3] | Mozambique, The Gambia, DRC, Tanzania, Kenya, Rwanda, Nigeria, Ghana, Uganda | 2005-2010 | 5425 | 527 (10%) | 2 (0-14) | 36% | 4211 | 4186 | 4786 |
| SMAC [4] | The Gambia, Kenya, Malawi, Ghana, Gabon | 2000-2005 | 26389 | 1136 (4%) | 2.2 (0-15) | 9% | 25845 | 25025 | 26199 |
| Lyke *et al.* [5] | Mali | 2000-2001 | 172 | 15 (9%) | 2.5 (0.2-10.4) | 21% | 166 | 166 | 172 |

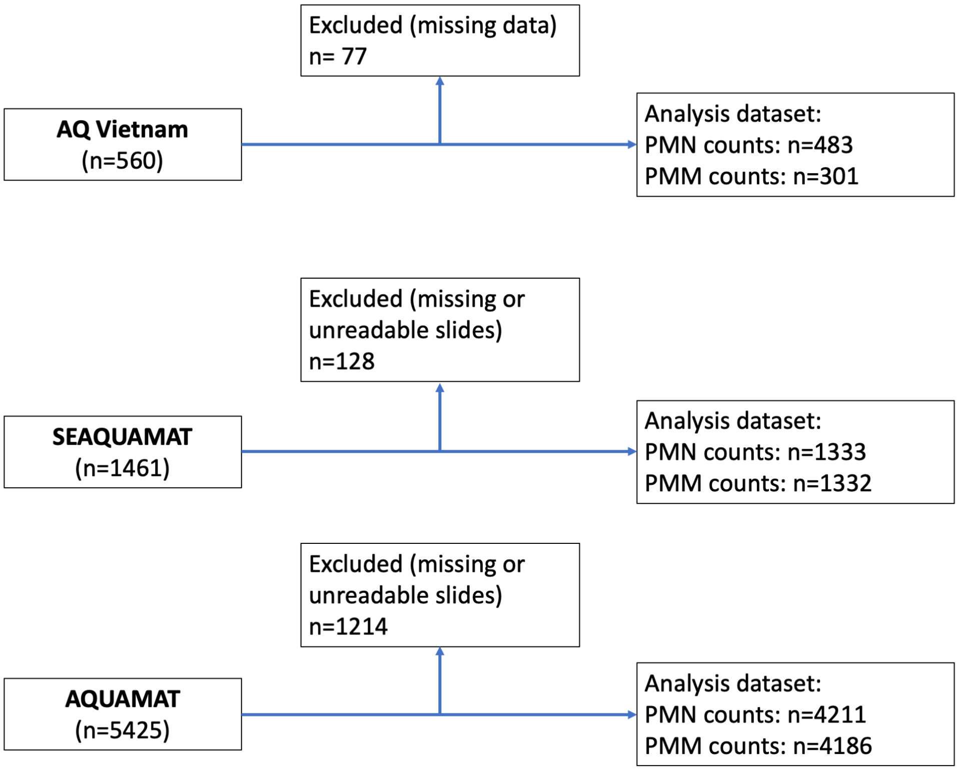


Figure S1: Flow diagram for inclusion and exclusion of data from the three randomised trials.

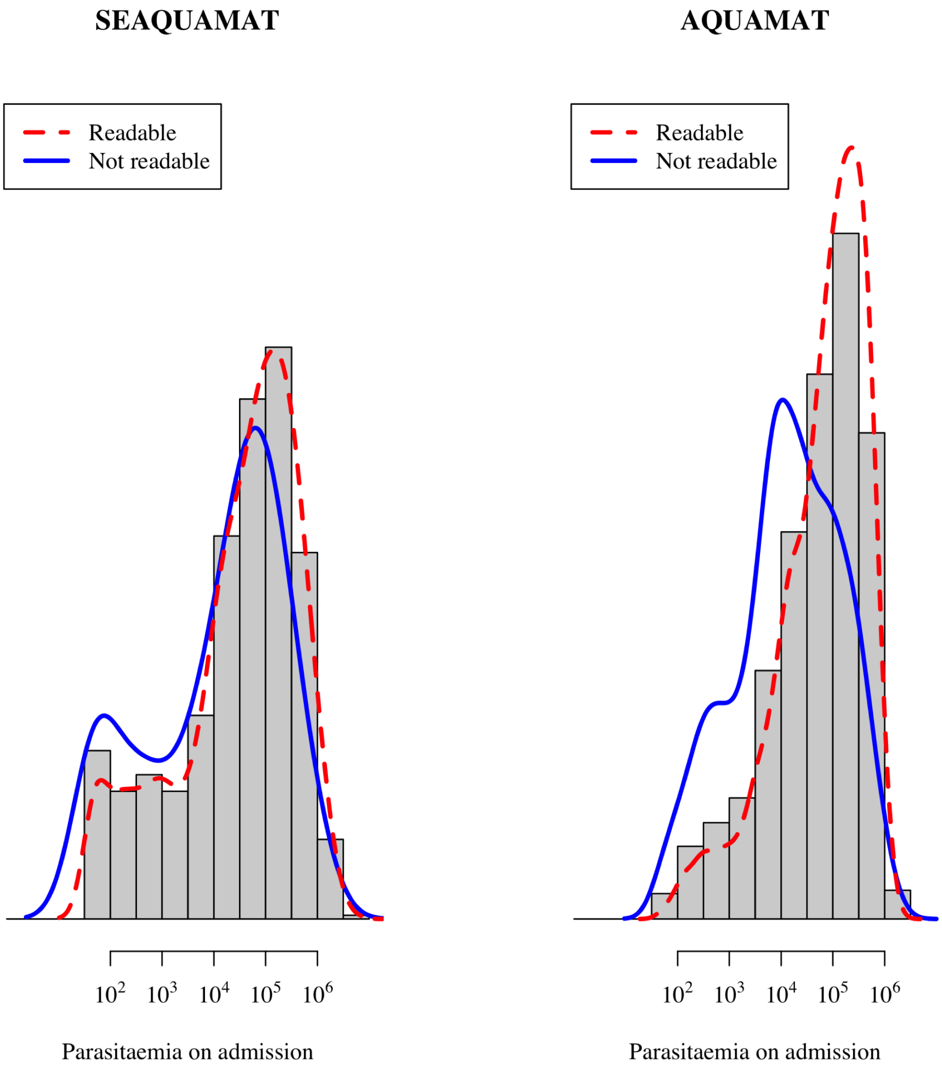


Figure S2: Relationship between malaria parasitaemia and data availability on pigment counts in the SEAQUAMAT and AQUAMAT randomised trials.

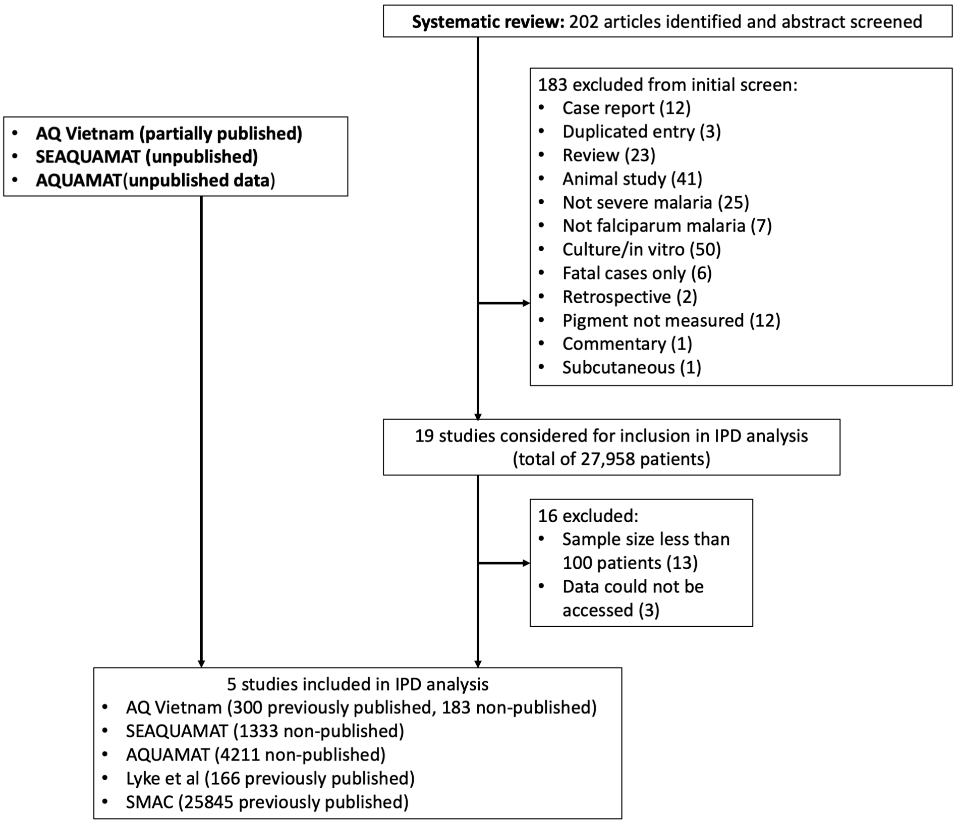


Figure S3: Flow diagram for inclusion and exclusion of data following the systematic review.

| Study | >5% pPMM Events | >5% pPMM Total | <5% pPMM Events | <5% pPMM Total | OR |
|---|---|---|---|---|---|
| AQ: Vietnam | 37 | 218 | 3 | 83 | 5.45 |
| SEAQUAMAT: Bangladesh | 32 | 125 | 82 | 285 | 0.85 |
| SEAQUAMAT: Myanmar | 40 | 197 | 42 | 356 | 1.90 |
| SEAQUAMAT: Indonesia | 7 | 23 | 17 | 212 | 5.02 |
| SEAQUAMAT: India | 20 | 49 | 12 | 85 | 4.20 |
| AQUAMAT: Mozambique | 18 | 185 | 18 | 209 | 1.14 |
| AQUAMAT: DRC | 12 | 293 | 3 | 87 | 1.20 |
| AQUAMAT: Ghana | 6 | 199 | 6 | 95 | 0.46 |
| AQUAMAT: Nigeria | 19 | 172 | 12 | 110 | 1.01 |
| AQUAMAT: Kenya | 28 | 208 | 8 | 93 | 1.65 |
| AQUAMAT: Tanzania | 112 | 742 | 59 | 465 | 1.22 |
| AQUAMAT: Rwanda | 6 | 100 | 6 | 105 | 1.05 |
| AQUAMAT: The Gambia | 30 | 267 | 21 | 219 | 1.19 |
| AQUAMAT: Uganda | 12 | 287 | 23 | 350 | 0.62 |
| SMAC: The Gambia | 152 | 1165 | 174 | 2278 | 1.81 |
| SMAC: Ghana | 84 | 1725 | 227 | 5071 | 1.09 |
| SMAC: Kenya | 43 | 1192 | 202 | 5604 | 1.00 |
| SMAC: Gabon (Lambarene) | 7 | 288 | 19 | 1523 | 1.97 |
| SMAC: Gabon (Libreville) | 11 | 203 | 38 | 703 | 1.00 |
| SMAC: Malawi | 22 | 492 | 113 | 4781 | 1.93 |
| Lyke et al: Mali | 10 | 109 | 5 | 57 | 1.05 |
| **Fixed effect model** | | **8239** | | **22771** | **1.35** |
| **Random effects model** | | | | | **1.37** |

Heterogeneity: $I^2$ = 60%, $\tau^2$ = 0.0809, $p$ = 2.45e-04

Test for overall effect (random effects): $z$ = 3.27 ($p$ = 1.1e-03)

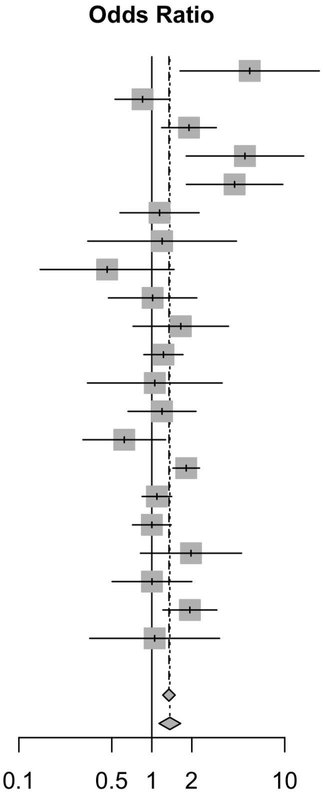


Figure S4: Meta-analysis of the prognostic value for mortality of >5% versus ≤ 5% pigment containing monocytes in 31,007 patients clinically diagnosed with severe falciparum malaria. We show the individual estimates broken down by country of enrolment for each study except for SMAC, Gabon where the distribution of pigment counts was substantially different between the two sites (Lambaréné and Libreville).

| Study | >5% pPMN Events | >5% pPMN Total | <5% pPMN Events | <5% pPMN Total | OR |
|---|---|---|---|---|---|
| AQ: Vietnam | 33 | 92 | 32 | 383 | 6.14 |
| SEAQUAMAT: Bangladesh | 54 | 160 | 50 | 220 | 1.73 |
| SEAQUAMAT: Myanmar | 52 | 185 | 22 | 269 | 4.39 |
| SEAQUAMAT: Indonesia | 9 | 25 | 11 | 138 | 6.49 |
| SEAQUAMAT: India | 16 | 52 | 15 | 78 | 1.87 |
| **Fixed effect model** | | **514** | | **1088** | **3.09** |
| **Random effects model** | | | | | **3.46** |

Heterogeneity: $I^2$ = 77%, $\tau^2$ = 0.3273, $p$ = 1.84e-03
Test for overall effect (random effects): $z$ = 4.15 ($p$ = 3.3e-05)

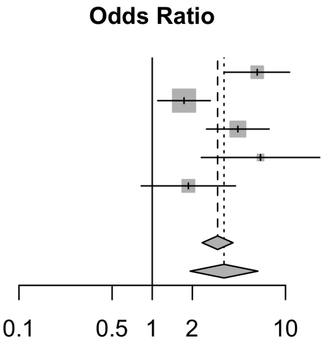


Figure S5: Pooled data set: prognostic value for mortality of >5% versus ≤ 5% pigment containing PMNs (neutrophils) in adults (defined as >15 years). None of the patients in the African studies were over 15 years of age.

| Study | >5% pPMN Events | >5% pPMN Total | <5% pPMN Events | <5% pPMN Total | OR |
|---|---|---|---|---|---|
| AQ: Vietnam | 0 | 1 | 1 | 7 | 1.44 |
| SEAQUAMAT: Bangladesh | 6 | 11 | 4 | 19 | 4.50 |
| SEAQUAMAT: Myanmar | 4 | 41 | 4 | 58 | 1.46 |
| SEAQUAMAT: Indonesia | 2 | 7 | 2 | 66 | 12.80 |
| SEAQUAMAT: India | 1 | 2 | 0 | 2 | 5.00 |
| AQUAMAT: Mozambique | 11 | 89 | 27 | 310 | 1.48 |
| AQUAMAT: DRC | 8 | 66 | 7 | 309 | 5.95 |
| AQUAMAT: Ghana | 4 | 101 | 8 | 194 | 0.96 |
| AQUAMAT: Nigeria | 13 | 66 | 18 | 218 | 2.73 |
| AQUAMAT: Kenya | 26 | 143 | 10 | 160 | 3.33 |
| AQUAMAT: Tanzania | 71 | 389 | 98 | 816 | 1.64 |
| AQUAMAT: Rwanda | 5 | 35 | 7 | 170 | 3.88 |
| AQUAMAT: The Gambia | 15 | 101 | 36 | 385 | 1.69 |
| AQUAMAT: Uganda | 6 | 90 | 30 | 547 | 1.23 |
| SMAC: The Gambia | 21 | 97 | 304 | 3342 | 2.76 |
| SMAC: Ghana | 73 | 686 | 238 | 6102 | 2.93 |
| SMAC: Kenya | 31 | 598 | 214 | 6195 | 1.53 |
| SMAC: Gabon (Lambarene) | 7 | 292 | 19 | 1514 | 1.93 |
| SMAC: Gabon (Libreville) | 13 | 109 | 75 | 1616 | 2.78 |
| SMAC: Malawi | 11 | 57 | 124 | 5198 | 9.79 |
| Lyke et al: Mali | 6 | 41 | 9 | 125 | 2.21 |
| **Fixed effect model** | | **3022** | | **27353** | **2.22** |
| **Random effects model** | | | | | **2.46** |

Heterogeneity: $I^2$ = 55%, $\tau^2$ = 0.1427, $p$ = 1.15e-03
Test for overall effect (random effects): $z$ = 7.10 ($p$ = 1.2e-12)

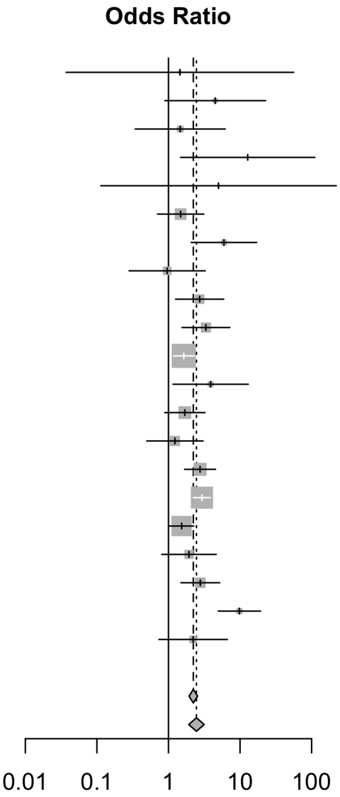


Figure S6: Pooled data set: prognostic value for mortality of >5% versus ≤ 5% pigment containing PMNs (neutrophils) in children (defined as ≤15 years).

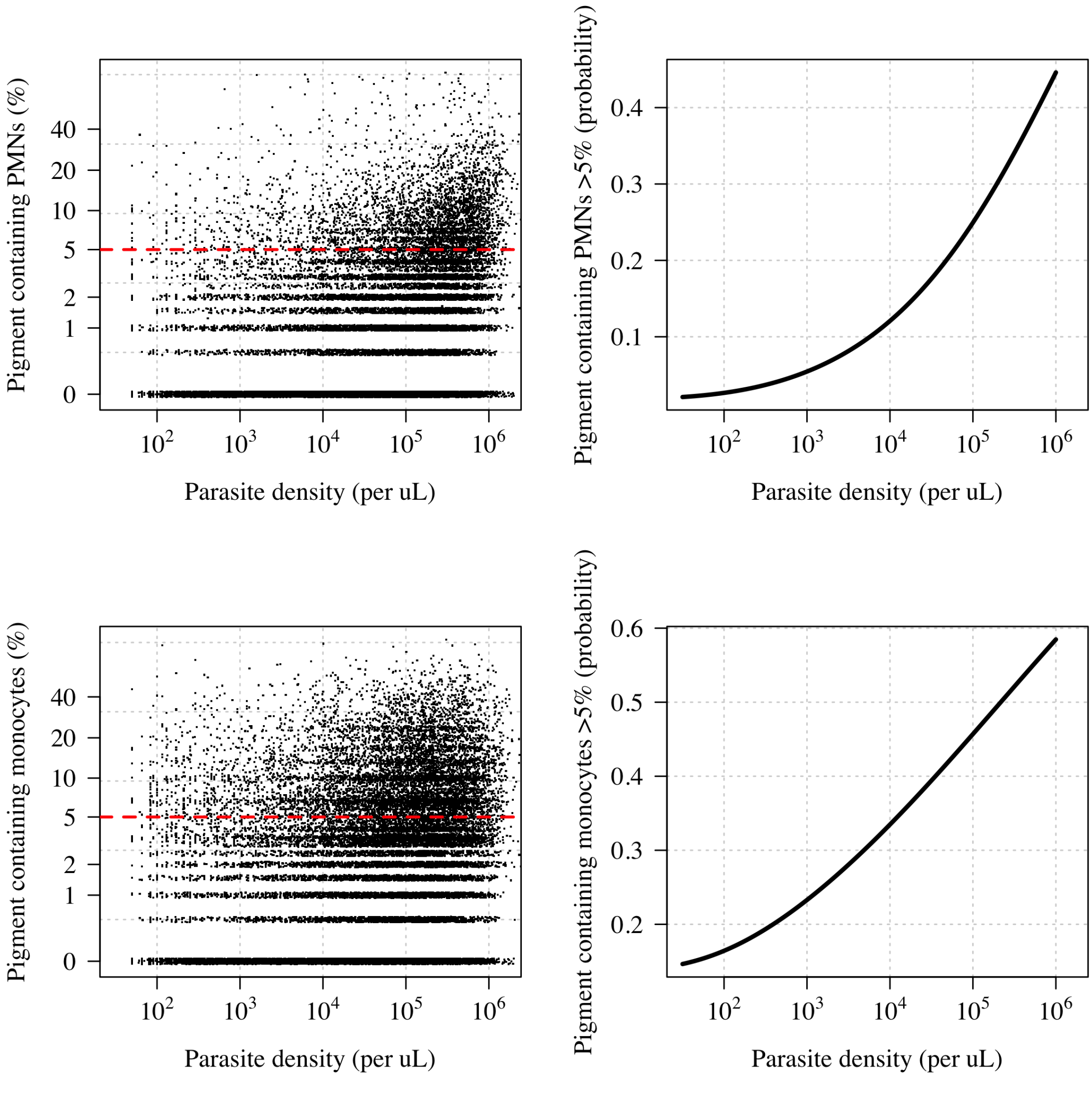


Figure S7: Relationship between parasite density and intraleukocytic malaria pigment (top: PMNs, bottom: monocytes). The left panels show scatterplots for the whole pooled data set; the right panels show the probability of having >5% pigment containing PMNs (neutrophils) or monocytes as a function of the peripheral blood malaria parasite density (fitted using a logistic regression model with nested random effects for site, country and study).

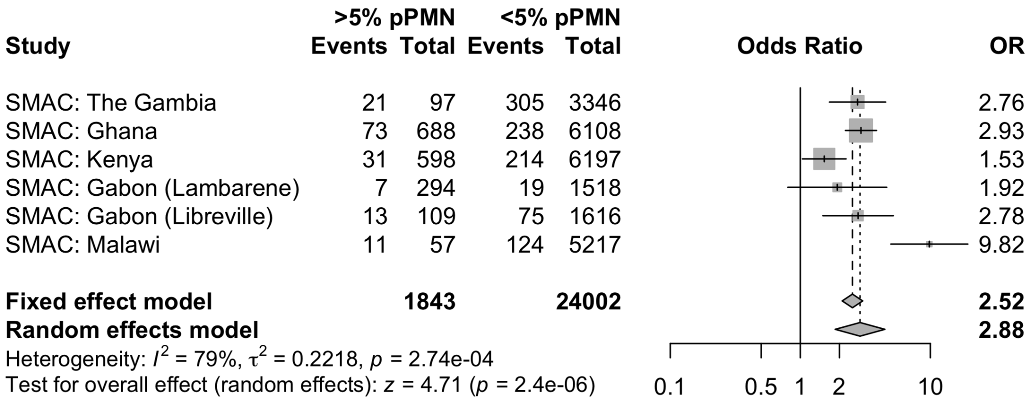


Figure S8: Prognostic value for mortality of $>5\%$ versus $\leq 5\%$ pigment containing PMNs (neutrophils) in patients from the SMAC study.

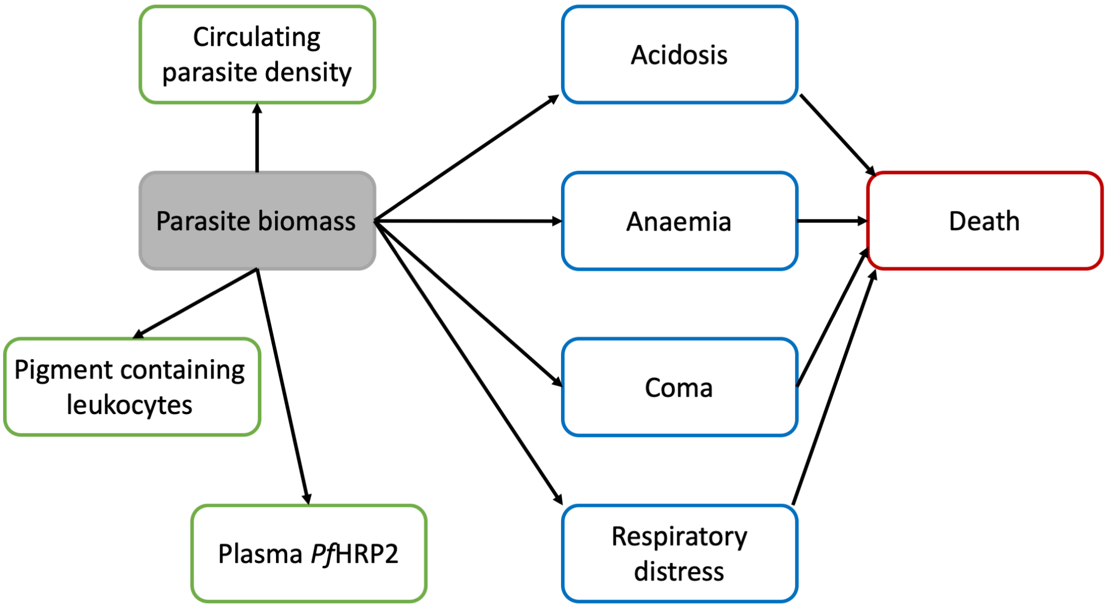


Figure S9: Proposed causal diagram for the relationship between intraleukocytic pigment counts, plasma *Pf*HRP2, parasite counts, and mortality in severe malaria. The main determinant of mortality is the parasite biomass which is unobservable (grey). The three exposure variables of interest (green) are proxy measurements of varying accuracy for the parasite biomass. The clinical variables in blue mediate the relationship between the parasite biomass and death (red).

### Systematic review

The systematic review identified 19 relevant publications which described clinical series in which pigment containing peripheral blood leukocytes had been counted. In most the the relationship to the severity of malaria (or death) had been evaluated [4–22]. The majority of these studies had very small sample sizes relative to the AQUAMAT and SEAQUAMAT randomised controlled trials (the median sample size was 67 for the number of severe malaria patients; some studies included uncomplicated malaria). We decided to contact study authors to obtain individual patient data for studies with at least 100 severe malaria patients. The data from Boeuf *et al* [16] could not be obtained as the principal investigator who had access to the data was deceased. The data from Luty *et al* could not be obtained as the authors could not find the original data. The data from Birhanu could not be obtained as the contact details were out of date and the corresponding author could not be contacted.

#### Previous reports

1. Phu *et al* (1995) described a prospective series of 300 consecutive adult Vietnamese patients with severe falciparum malaria (AQ Vietnam trial). The 40 who died had significantly higher proportions of malaria pigment-containing neutrophils on admission (mean=7.7%, standard deviation (SD)=5.9%) and pigment-containing monocytes (mean=8.6%, SD=5.9%) than did survivors (mean 3.2%, SD=4.1% and mean 4.8% (SD 4.6%) respectively) (P<0.0001). A count of peripheral neutrophils containing visible pigment >5% predicted a fatal outcome with 73% sensitivity and 77% specificity (relative risk 6.2, 95% confidence interval (CI) 3.2-11.8) compared to 60% sensitivity and 57% specificity for parasitaemia >100,000/uL (relative risk 1.8, 95% CI 1.0-3.3).

2. Metzger *et al* (1995) reported a series from Gabon comprising 42 children with severe malaria, 31 children with "mild" malaria, and 31 adults with "mild" malaria. More pigment-containing monocytes and pigment-containing neutrophils were found in severe malaria. There was one death (11% pigment containing neutrophils). The authors concluded that the elevation in pigment-containing monocytes was more informative.

3. Amodu *et al* (1998) described a series of 146 Nigerian children aged between 6 months and 14 years and divided them in four categories, of whom 43 had severe malaria. The median proportion of pigment-containing neutrophils showed a clear rise across the spectrum from no malaria (2.0%), asymptomatic malaria (6.5%), mild malaria (9.0%) and cerebral malaria (27.0%) (P < 0.0001). The proportion of pigment-containing monocytes did not differ significantly between the mild malaria, asymptomatic malaria and no malaria groups but the cerebral malaria group had a higher median value than the other three groups.

4. Luty *et al* (2000) conducted a case-control comparison of 100 patients with severe malaria (cases) and 100 patients with mild malaria (controls). The proportions of pigment-containing monocytes and pigment-containing neutrophils on admission were positively correlated with parasitaemia and were 4- and 18-fold-larger respectively in severe compared with mild malaria

5. Lyke *et al* (2003) studied 172 Malian children with severe malaria and 172 controls with uncomplicated malaria. The presence of polymorphonuclear cell (PMN) and monocyte (PMM) pigment was strongly associated with severe disease compared with uncomplicated malaria. Total PMN pigment burden in children with severe malaria was higher in those with cerebral manifestations and with combined cerebral manifestations and severe anaemia (haemoglobin <5 g/dL) but was not associated with hyperparasitaemia (> 500,000 asexual forms/mm3). Additionally, the count of pigment containing PMNs was associated with a fatal outcome in patients with severe malaria.

6. Ochola *et al* (2005) studied prospectively 22 Kenyan children with severe malaria. Peripheral parasitaemia, plasma levels of sTNF-R75 and circulating parasite DNA correlated better with estimated sequestered parasite load than pigment containing leukocytes.

7. Mujuzi *et al* (2006) conducted a case-control study of infants with severe malaria in Uganda; 99 with severe malaria and 96 with uncomplicated malaria. Higher proportions of pigment containing monocytes were associated with anaemia.

8. Casals-Pascual *et al* (2006) studied 26 Kenyan children with severe malarial anaemia. Haemozoin-containing neutrophils were strongly associated with plasma lactate levels (a well characterized prognostic measure in severe malaria), independently of parasite density ($r = 0.60$, $P = 0.003$).

9. Hanscheid *et al* (2008) studied 152 patients in Gabon with falciparum malaria. Compared with the 104 children with uncomplicated malaria the 48 children with severe malaria had higher proportions of pigment-containing neutrophils but not monocytes.

10. Kremsner *et al* (2009) analysed pigment containing leukocytes in the admission thick blood smears of 26,296 children hospitalized with P. falciparum and studied in the Severe Malaria in African Children network. They found that at all but one of the study sites the mortality for patients with >5 pigment containing granulocytes per 200 white blood cells was higher than in subjects with no pigment containing granulocytes. However, they described adjusted odds ratios for associations between mortality and pigment containing leukocytes, estimated through logistic regression and which included other established markers of severe malaria, as "moderate to nonexistent in most sites". They concluded that "levels of pigmented cells were not useful predictors of outcome across Africa".

11. Novelli *et al* (2010) studied 355 Kenya children falciparum malaria, into three groups: uncomplicated malaria (Hb >11.0 g/dL, $n$=23); anaemic (Hb 6-10.9g/dL, $n$=200), and severe malarial anaemia (Hb < 6.0 d/dL, $n$=132). In a multiple logistic regression analysis pigment containing monocytes were associated independently with severe malarial anaemia. There were no deaths in the severe malaria group.

12. Fendel *et al* (2010) studied a series of children with severe malarial anaemia in Gabon. The proportions of pigment containing monocytes were higher in the 39 children with severe anaemia compared with values in the 52 with mild malaria.

13. Boeuf *et al* (2012) conducted a prospective study of Ghanaian children with falciparum mlaria; 144 had cerebral malaria, 108 severe anaemia and 80 had uncomplicated malaria. The proportions of pigment-containing monocytes were higher in cerebral malaria than in severe anaemia, and both were higher than in uncomplicated malaria.

14. Abugri *et al* (2014) studied 100 children with falciparum malaria in Ghana. The proportions of pigment-containing monocytes were higher in severe malaria ($n$=32) than they were in uncomplicated malaria ($n$=68).

15. Kempaiah (2016) studied 144 children with falciparum malaria, stratified into two clinical categories: non-SMA (Hb >5.0 g/dL, $n$=91) and SMA (Hb <5.0 g/dL, $n$=53). They did a whole of complicated lab stuff and made some conclusions about things that probably have zero clinical utility. Their best result was to show that people with severe anaemia (Hb<5) had a lower haemoglobin than people without severe anaemia (Hb>5) with an incredible p-value <0.0001. A real cracker.

16. Mandala (2016) enrolled children presenting with cerebral malaria ($n$=36), severe malarial anaemia ($n$=42) or uncomplicated malaria ($n$=66), and healthy aparasitemic children ($n$=52) in Blantyre, Malawi. They observed that in all malaria groups, especially the severe malarial anaemia group, a greater proportion of monocytes contained haemozoin pigment compared to controls.

17. Birhanu *et al* (2017) studied children hospitalized with malaria in Ethiopia, 283 of whom had *P. falciparum* infections, 57 had *P. vivax* and 37 were mixed. Of these 102 children had severe malaria. Both elevated pigment-containing monocytes and pigment-containing neutrophils were associated independently with severe malaria (AOR = 6.26, 95% CI: 2.14–14.29 and AOR = 7.93, 95% CI: 3.09–16.86 respectively).

18. Salih *et al* (2018) conducted a case control study in Sudanese children; 67 with severe falciparum malaria, 63 with uncomplicated falciparum malaria and 50 healthy children. Haemozoin containing monocytes were more common in severe malaria than in uncomplicated malaria.

| | Patient selection (risk of bias) | Microscopy evaluation (risk of bias) | Outcome evaluation (risk of bias) | Patient selection (applicability) | Outcome evaluation (applicability) |
|---|---|---|---|---|---|
| AQ Vietnam [1] | LOW | LOW | LOW | LOW | HIGH |
| SEAQUAMAT [2] | LOW | LOW | LOW | LOW | HIGH |
| AQUAMAT [3] | LOW | LOW | LOW | MODERATE | HIGH |
| Lyke et al [5] | LOW | LOW | LOW | MODERATE | HIGH |
| SMAC [4] | LOW | LOW | LOW | MODERATE | HIGH |

Table S2: Assessment of the risk of bias and applicability of the included studies using a modified QUADAS-2 tool [23].

19. Achieng *et al* (2019) compared 53 parasitaemic Kenyan children with severe malaria anaemia SMA (Hb<5.0g/dL) and 69 parasitaemic children with higher haemoglobin values. Counts of pigment-containing monocytes and pigment-containing neutrophils were higher in children severe malaria anaemia (P=.028 and P=.079, respectively).

The study of Davenport *et al* (2010) is not included in our list as this concentrated on the impact of HIV infection on clinical severity and longer term prognosis. This was a study of 542 Kenyan children aged 3-36 months with falciparum malaria. HIV positive children had significantly more malarial pigment-containing neutrophils, monocytosis, and were more likely to be severely anaemic increased (Hb $< 6.0$ g/dL), and were nearly ten times more likely to die within 3 months of enrollment. Increased proportions of pigment-containing monocytes but not pigment-containing neutrophils were associated significantly with severe malarial anaemia. Patients with any density of *P. falciparum* were included in the study. None of the children had cerebral malaria and it is unclear how many would have met the WHO definition for severe malarial anaemia.

### Data collection from included studies

For all studies that passed screening and had more than 100 patients with severe malaria, we contacted the corresponding author using available emails addresses found online. For the SMAC study, the data had been deposited open access (https://dataverse.harvard.edu/dataset.xhtml?persistentId=doi:10.7910/DVN/OCTWUJ) but we checked our findings with the corresponding author. One author could not be contacted (Birhanu et al).

Data requested for each study included all intraleukocytic pigment counts (number of cells counted and number containing pigment), the parasite count at baseline, in-hospital mortality outcome, and any available baseline clinical or laboratory variables. For each included dataset we plotted all relevant data to check integrity and data quality. R scripts used to clean the SMAC study data are available on the linked github repository (https://github.com/jwatowatson/MalariaPigmentPrognosis).

### Risk of bias and applicability using a modified QUADAS-2 criteria tool

In all five studies, the patients were hospital cohorts (no risk of bias in how they were selected). In all studies microscopy was done blinded to patient allocation (randomised trials) and outcome. For all trials the outcome was in-hospital mortality (very low risk of bias).
